## Supplementary material for "Nutritional Assessment and Associated Factors in Children with Congenital Heart Disease - Ethiopia": abstract

**Covering Letter**

Congenital heart disease is one of the major congenital anomalies occurring worldwide with a prevalence close to 1 in 1000 live births. As the fertility rate is high in developing countries this problem is seen at higher rate in these countries.

Many literatures described the correlation between congenital heart diseases and malnutrition. This problem tends to be more in developing countries because these patients do not get the necessary intervention at appropriate time. Likewise, malnutrition itself is quite common in these countries because of other reasons.

If this manuscript is published it will be helpful for the health professional especially for undergraduate medical students, pediatric residents and pediatricians to let them know the correlation between congenital heart diseases and malnutrition and also other contributing factors for this cooccurrence. It also describes the need for early intervention in these patients.

We are also currently conducting a research as a continuation of this one as we follow pediatric patients with congenital heart disease and malnutrition for whom intervention was done. By doing which we will try to see the short- and long-term impact of intervention on malnutrition.

We would like to say thank you in advance for your consideration and your valuable time spent in reviewing this manuscript. We are glad to respond for any clarification or supplements you inquire with this regard.
